# The risk and risk factors of chikungunya virus infection and rheumatological sequelae in a cohort of U.S. Military Health System beneficiaries: implications for the vaccine era

**DOI:** 10.1101/2023.11.22.23298875

**Authors:** SD Pollett, H-C Hsieh, D Lu, M Grance, G Nowak, C Lanteri, D Tribble, TH Burgess

## Abstract

**Background:** Understanding the risk of chikungunya virus (CHIKV) infection and rheumatic sequelae across populations, including travelers and the military, is critical. We leveraged the electronic medical records of about 9.5 million U.S. Military Health System (MHS) beneficiaries to identify the risk of post-CHIKV rheumatic sequelae.

**Methodology/Principal Findings:** MHS beneficiary CHIKV infections diagnosed 2014–2018 were identified from the Disease Reporting System internet, TRICARE Encounter Data Non-Institutional, and Comprehensive Ambulatory/Professional Encounter Record systems. Non-CHIKV controls were matched (1:4) by age, gender, beneficiary status, and encounter date. The frequency of comorbidities and incident rheumatic diagnoses through 2020 were derived from International Classification of Diseases codes and compared between cases and controls. Logistic regression models estimated the association of CHIKV infection with rheumatic sequelae and risk factors for post-CHIKV sequelae. 195 CHIKV cases were diagnosed between July 2014 and December 2018. The mean age was 42 years, and 43.6% were active duty. 63/195 (32.3%) of CHIKV cases had an incident rheumatic diagnosis, including arthralgia, polyarthritis, polymyalgia rheumatica, and/or rheumatoid arthritis, compared to 156/780 (20.0%) of controls (p < 0.001). CHIKV infection remained associated with rheumatic sequelae (aOR = 1.911, p = 0.002) after adjusting for prior rheumatic disease and demography. Those with rheumatic CHIKV sequelae had a median 7 healthcare encounters (IQR 3–15). Among CHIKV infections, we found no association between post-CHIKV rheumatic sequelae and demography, service characteristics, or comorbidities.

**Conclusions/Significance:** CHIKV infection is uncommon but associated with rheumatic sequelae among MHS beneficiaries, with substantial healthcare requirements in a proportion of cases with such sequelae. No demographic, clinical, or occupational variables were associated with post-CHIKV rheumatic sequelae, suggesting that prediction of these complications is challenging in MHS beneficiaries. These findings are important context for future CHIKV vaccine decision making in this and other populations.

**Author summary:** We examined U.S. Military Health System (MHS) electronic medical records during to identify the likelihood of rheumatic complications after chikungunya virus (CHIKV) infection. Overall, CHIKV infections were rare in the MHS, with 195 cases found in the records between 2014 and 2018 (a period which encompassed the peak of the CHIKV epidemic in the Americas). Of these, about 32% received a rheumatic diagnosis after infection, including arthralgia, polyarthritis, polymyalgia rheumatica, and rheumatoid arthritis. Patients who had a rheumatic diagnosis had on average 7 healthcare encounters for their post-CHIKV rheumatic complication, and a quarter had more than 15 healthcare encounters. We did not find any demographic, clinical, or occupational characteristics associated with developing rheumatic complications after CHIKV, suggesting that predicting rheumatic complications from CHIKV may be challenging in MHS beneficiaries. These findings may provide important context for decisions about implementing an approved chikungunya vaccine to military servicemembers and other MHS beneficiaries.

## Introduction

Chikungunya virus (CHIKV) is a mosquito-borne alphavirus belonging to the *Togaviridae* family [1]. The ecology of CHIKV is characterized by a sylvatic cycle involving non-human primates and arboreal *Aedes* species and urban human-to-human transmission cycles involving the *A. aegypti* and *A. albopictus* vectors [1]. While CHIKV is defined by a single serotype, multiple genotypes have evolved with global geographic expansion including the Asian and descendant Asian-Caribbean lineages, and East/Central/South African and Indian Ocean lineages [1–3].

The 2013–2015 neotropical epidemic in the Americas involved over 1.6 million suspected or confirmed CHIKV cases [4]. While cases rapidly declined after the initial neotropical pandemic, there is an ongoing substantive CHIKV burden in the Americas in addition to ongoing circulation in Africa [5, 6] and Asia [7, 8]. Recently, the Pan American Health Organization issued an alert indicating the geographic range of CHIKV has expanded in the Americas, with notable resurgence in certain regions of Latin America [9–11].

Acute CHIKV infection illness is characterized by a high symptomatic-to-asymptomatic ratio, with clinical features including an abrupt onset of high fever, polyarthralgia, backache, headache, and fatigue [12]. Atypical acute CHIKV illness manifestations (e.g., acute encephalitis) are rare, potentially severe, and more often occur in the extremes of age and in those with underlying comorbidities [12–17]. Diagnosis of CHIKV infection relies on blood polymerase chain reaction (PCR) within the first week of illness and serology thereafter [18]. Management of acute CHIKV is generally supportive, with fluid replacement, analgesia and avoidance of non-steroidal anti-inflammatory drugs until dengue has been excluded as a differential diagnosis [19]. There are no current vaccines or other preventive medical countermeasures available for CHIKV aside from avoidance of vectors in endemic transmission regions [19]. Advanced vaccine development has been challenging due to the unpredictable nature of CHIKV outbreaks [20]. However, CHIKV vaccine U.S. Food and Drug Administration licensure may soon occur using a serological correlate of protection [21].

The rheumatological manifestations of CHIKV may be categorized by timing and clinical phenotype [22]. Acute CHIKV arthralgia is frequently reported and mostly occurs in mostly peripheral joints with a symmetrical pattern, although large joints can be affected [22]. Joint swelling and synovitis often accompanies this acute polyarthralgia; functional impairment during this acute phase is common due debilitating pain [22]. Chronic rheumatological manifestations typically involve the joints affected in acute CHIKV arthralgia [22]. Estimates on the frequency of persistent post-acute manifestations (predominantly arthralgia or arthritis) vary significantly, with metanalysis estimates as high as 43% at 3 months, 21% at 12 months, and 14% at 18 months after initial CHIKV illness onset [22–24]. This heterogeneity is believed to reflect differences in study populations and methodologies. Pathogenesis studies have indicated that persistent host inflammation, rather than persistent synovial viral replication, plays a key role in post-CHIKV arthralgia [25, 26]. There is currently a wide variation in clinical management for chronic post-CHIKV sequelae [15] but post-acute CHIKV arthropathy is severe enough in some to warrant the use of biological or disease-modifying antirheumatic drugs [27].

While those residing in CHIKV endemic regions experience the highest burden of this arbovirus, travelers and the deploying military are also significant stakeholders for understanding the risk of CHIKV infection and post-CHIKV sequelae. There is limited published data on the burden of CHIKV in U.S. Military Health System (MHS) beneficiaries [28, 29] and none define the risk and risk factors of post-acute CHIKV sequelae in this population. Understanding the risk and risk factors of CHIKV and CHIKV sequelae is important for clinical prognostication and may support cost-effectiveness analyses for vaccine policy with potential future licensed vaccines; such findings may be generalizable to traveler populations also.

The objectives in this study were to describe to characterize incident CHIKV cases that were diagnosed in the MHS between 2014–2018 and to further describe the risk and risk factors of acute and chronic post-CHIKV rheumatological sequelae in such CHIKV cases, compared to matched controls. Here, we leveraged a virtual cohort derived from over 9.5 million MHS beneficiaries, with follow-up through 2020.

## Methods

### Population, setting and identification of CHIKV cases and matched controls

The MHS serves approximately 9.5 million beneficiaries, including active duty servicemembers, retirees and dependents. This includes care delivered within the United States, U.S. Territories (including Puerto Rico, Guam, and the U.S. Virgin Islands), and in beneficiaries located other countries.

For this study, we identified deduplicated unique CHIKV cases in MHS beneficiaries via multiple data streams from 2005 through 2018, including: (i) clinical microbiology lab (serology and PCR) results from Disease Reporting System internet (DRSi)/EpiData Center, (ii) International Classification of Diseases, 10th Revision (ICD-10) code A92.0 CHIKV disease entries from TRICARE Encounter Data Non-Institutional (TEDNI), and (iii) ICD-10 code A92.0 CHIKV disease from Comprehensive Ambulatory/Professional Encounter Record (CAPER). The treatment locations of cases were inferred by clinical provider ZIP code. To estimate the risk of post-CHIKV rheumatological sequalae, CHIKV negative MHS beneficiary controls were matched (1:4) with cases by age group, gender, date of healthcare encounter (+/- 1 month), and beneficiary status at the time of matching healthcare encounter. Controls were randomly selected within these matching strata.

### Demographic, post-CHIKV diagnosis and comorbidity data abstraction from the electronic medical record

Demographic ICD-9 and ICD-10 code data abstracted from cases and controls (using Armed Forces Health Longitudinal Technology Application (AHLTA) and the Composite Health Care System (CHCS). The sequelae of interest were any rheumatologic disorders (See S1 Table for ICD-9 and ICD-10 codes). Co-morbidity was defined as a condition/disorder (including rheumatological) which pre-existed before the time of the first CHIKV infection diagnosis (or first matched healthcare encounter of the time-matched control. We defined post-CHIKV rheumatological complications as new post-CHIKV rheumatological diagnoses with a first onset at or after the first CHIKV diagnosis (or the healthcare encounter used for matching for controls without CHIKV). When a pre-CHIKV rheumatological and post-CHIKV rheumatological diagnosis mapped to the same ICD code, we further adjudicated a pre-existing rheumatological diagnoses from a new post-CHIKV complication using DX_DESC-level diagnosis coding (Agency for Healthcare Research and Quality Clinical Classifications Software system code). A similar approach was taken for controls without CHIKV.

### Statistical analysis

Demographic, clinical, and occupational characteristics of cases and controls were summarized. The frequency of rheumatological sequelae were compared in cases and controls. Conditional logistic regression (on age and sex matched cases and controls) were used to estimate the independent association between CHIKV infection and short-term (<3 months) versus long-term (≥3 months) complications, adjusting for prior comorbidities (including pre-existing rheumatological comorbidities). Unadjusted and adjusted odds ratios were presented. Finally, among those with CHIKV infections, we performed multinomial regression for any to determine risk factors for any rheumatologic related sequelae. For each multinomial regression analysis, there were 3 comparisons: participants with short-term (<3 months), long-term (≥3 months) and no sequelae.

All p values were two-sided, with an alpha significance threshold of 0.05. Analyses were conducted using SAS software (version 9.4; SAS).

### Ethical considerations

This protocol was reviewed and approved for execution by the IRB of the Uniformed Services University of the Health Sciences, Bethesda, Maryland, United States of America.

## Results

We identified 195 unique CHIKV cases diagnosed in the MHS between July 2014 and December 2018 (S1 Fig). The most frequent locations of clinical encounters were in Puerto Rico, Texas, and California, but these reflect provider ZIP codes of diagnoses (including potential diagnoses made after deployments or other beneficiary travel) and do not necessarily reflect location of where cases acquired CHIKV infections (Fig 1). Our finding of the majority of cases being diagnosed in Puerto Rico agreed with prior, smaller studies in MHS beneficiaries [29, 30]. Time series of all MHS CHIKV cases (by month) indicated the peak of MHS CHIKV cases occurring in late 2014 during the peak year of the Americas CHIKV epidemic.

**Fig 1.**
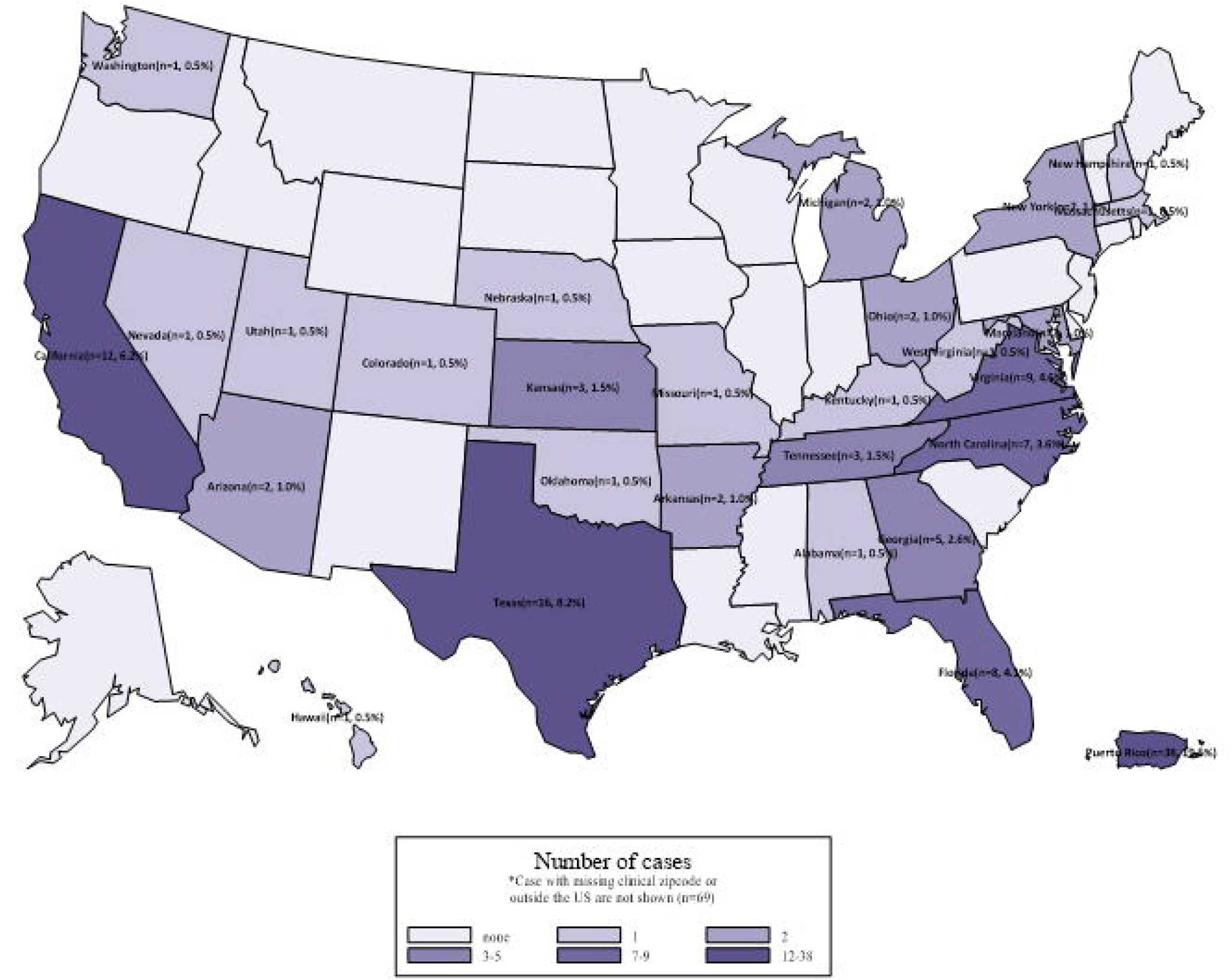
Location of CHIKV cases by U.S. state or territory.

The median age of CHIKV cases was 42 years (IQR 31–54 years), and 43.6% were active duty servicemembers (Table 1). Compared to age, gender, beneficiary status, and healthcare encounter-matched controls, cases differed from controls with respect to race (p < 0.001). Among active duty CHIKV cases, we noted that Army service and enlisted rank were most frequent; the frequency of these categories were more common than matched controls (p < 0.001; Table 1). CHIKV cases had a higher frequency of all measured comorbidities compared to age, sex, and beneficiary-matched controls (p < 0.001; Table 2).

**Table 1.**
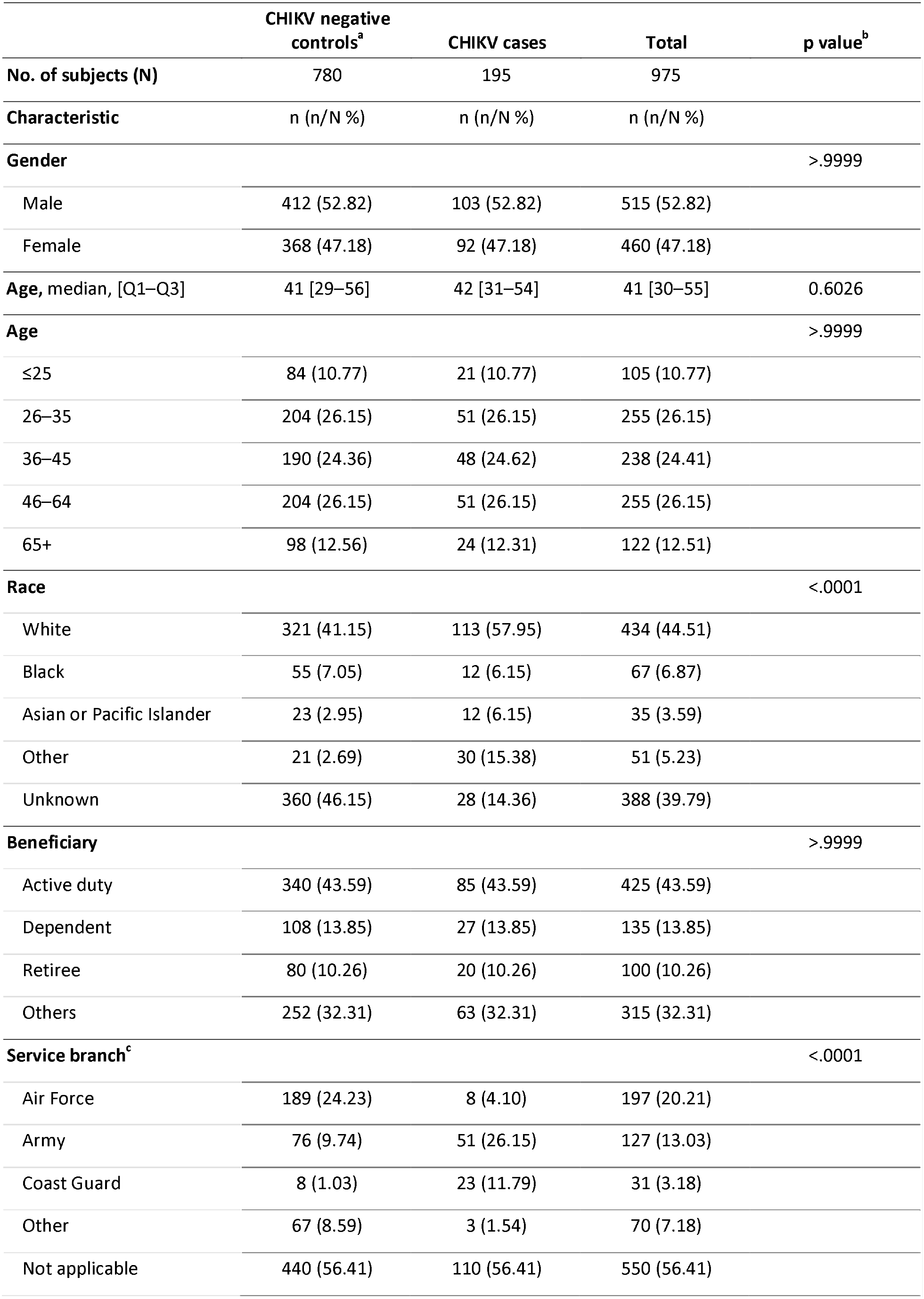
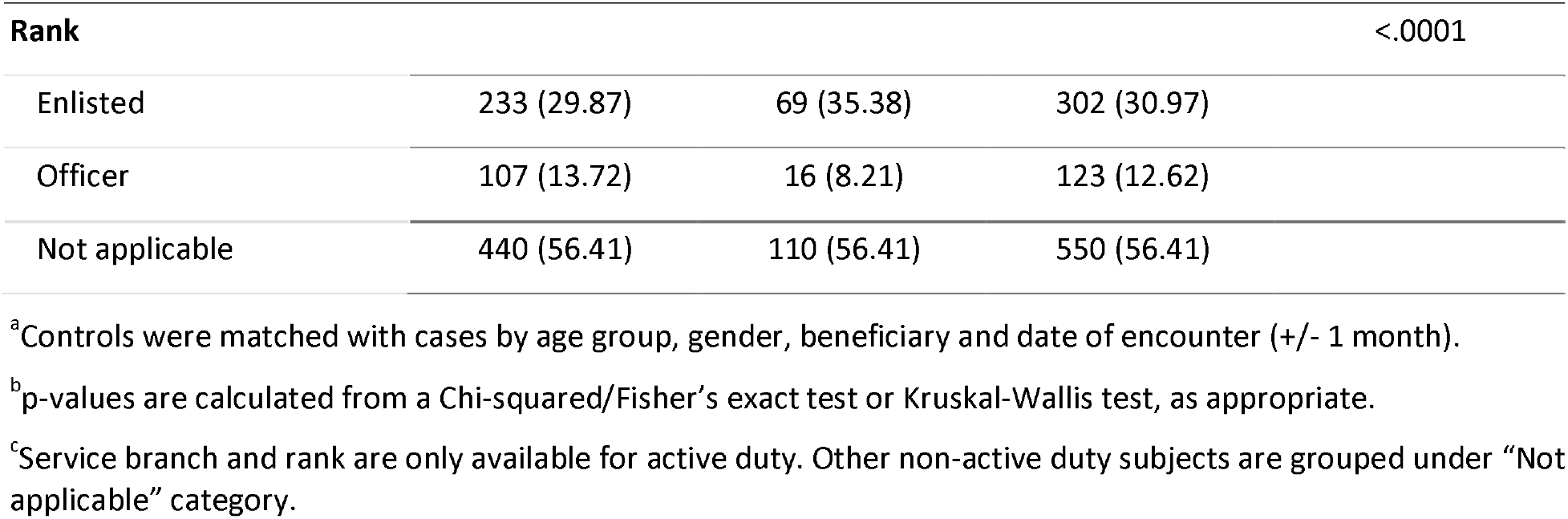
Demographic and professional characteristics in CHIKV cases and controls.

**Table 2.** Comorbidities in CHIKV cases and controls.

|  | CHIK- | CHIK+ | Total | p value<sup>a</sup> |
| --- | --- | --- | --- | --- |
| **Total # of subjects** | 780 | 195 | 975 |  |
| **Comorbidities (ICD diagnoses before time of matching)** |  |  |  |  |
| None | 461 (59.10) | 34 (17.44) | 495 (50.77) | <.0001 |
| Cancer | 30 (3.85) | 22 (11.28) | 52 (5.33) | <.0001 |
| Cardiovascular disease | 196 (25.13) | 95 (48.72) | 291 (29.85) | <.0001 |
| Chronic respiratory disease | 91 (11.67) | 58 (29.74) | 149 (15.28) | <.0001 |
| Mental health condition | 117 (15.00) | 72 (36.92) | 189 (19.38) | <.0001 |
| Osteoporosis | 26 (3.33) | 19 (9.74) | 45 (4.62) | 0.0001 |
| Upper gastrointestinal disease | 103 (13.21) | 75 (38.46) | 178 (18.26) | <.0001 |
| Neurologic disease | 122 (15.64) | 80 (41.03) | 202 (20.72) | <.0001 |
| Rheumatologic disease | 117 (15.00) | 84 (43.08) | 201 (20.62) | <.0001 |
| Osteoarthritis | 92 (11.79) | 55 (28.21) | 147 (15.08) | <.0001 |
| Other non-traumatic joint disorders | 29 (3.72) | 32 (16.41) | 61 (6.26) | <.0001 |
| Rheumatoid arthritis and related disease | 12 (1.54) | 17 (8.72) | 29 (2.97) | <.0001 |
| Spondylosis; disc disorders; other back problems | 51 (6.54) | 37 (18.97) | 88 (9.03) | <.0001 |

<sup>a</sup>p values were calculated from a Chi-squared/Fisher's exact test.

We noted 63/195 (32.3%) of CHIKV cases had an incident post-CHIKV rheumatological diagnosis, compared to 156/780 (20.0%) of controls (p = 0.0002) (Table 3). Osteoarthritis, rheumatoid arthritis and related diagnoses (including inflammatory polyarthropathy), and non-traumatic joint disorder associated categories (including unspecified arthropathy, polyarthralgia and polyarthritis) were noted with statistically significant higher frequency in post-CHIKV cases versus controls (Table 3, S2 Table).

**Table 3.**
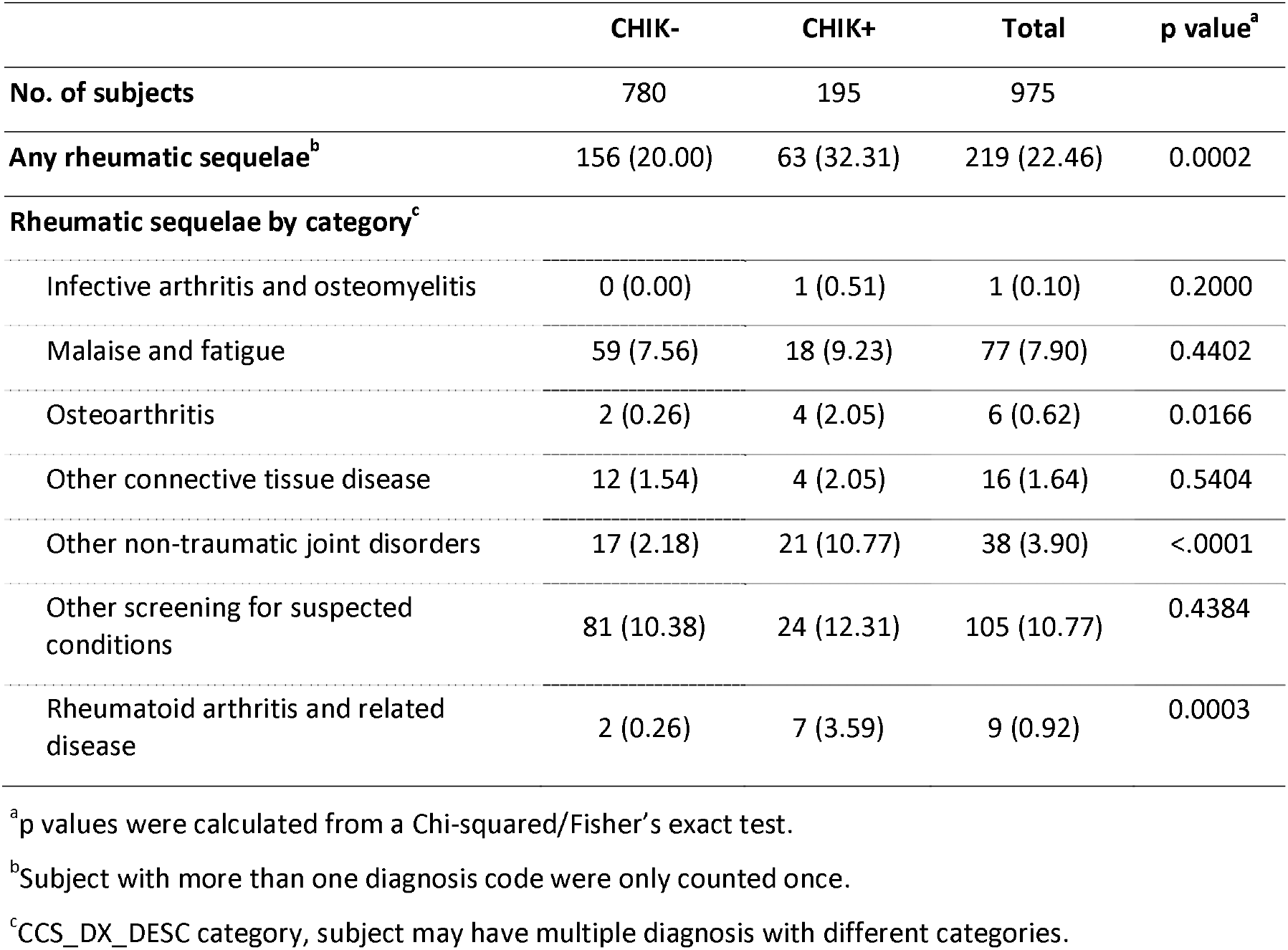
Rheumatological sequelae in CHIKV cases and controls.

Compared to controls, CHIKV infection was associated with post-infection rheumatological diagnoses (aOR = 1.91, 95% CI 1.272–2.870, p = 0.0018) after adjusting for prior the presence of any prior pre-CHIKV rheumatological diagnoses, beneficiary status, age, sex, and race (Table 4). The magnitude of this effect size was larger when the outcome was stratified into short-term (≤3 month) post-CHIKV rheumatological diagnoses versus long-term (diagnosis recurred beyond 3 months post-CHIKV infection) rheumatic sequelae (aOR = 6.41, p < 0.001 short-term sequelae; aOR = 1.43, p = 0.09 long-term sequelae, respectively) (Table 5). Those with post-CHIKV rheumatological sequelae had a median 7 healthcare encounters for rheumatological diagnoses (IQR 3–15, range 1–33) through the period of observation.

**Table 4.** Unadjusted and adjusted odds ratio of CHIKV infection on rheumatic-related sequelae (n = 975).

|  | Unadjusted |  |  |  | Adjusted<sup>a</sup> |  |  |  |
| --- | --- | --- | --- | --- | --- | --- | --- | --- |
|  | OR | 95 % CI |  | p value | OR | 95% CI |  | p value |
| CHIK positive | 1.909 | 1.348 | 2.703 | 0.0003 | 1.911 | 1.272 | 2.870 | 0.0018 |
| Rheumatic related comorbidity | 1.609 | 1.134 | 2.283 | 0.0078 | 1.137 | 0.763 | 1.695 | 0.5283 |
| Age | 1.011 | 1.002 | 1.019 | 0.0107 | 1.012 | 1.002 | 1.022 | 0.0181 |
| Male | 0.984 | 0.728 | 1.330 | 0.9171 | 0.918 | 0.620 | 1.358 | 0.6669 |
| Race (ref: White) |  |  |  |  |  |  |  |  |
| Black | 1.062 | 0.580 | 1.944 | 0.8464 | 1.178 | 0.633 | 2.192 | 0.6055 |
| Asian or Pacific Islander | 0.846 | 0.359 | 1.995 | 0.7024 | 0.758 | 0.314 | 1.830 | 0.3551 |
| Other | 0.960 | 0.698 | 1.319 | 0.8001 | 1.294 | 0.846 | 1.977 | 0.2307 |
| Beneficiary (ref: active duty) |  |  |  |  |  |  |  |  |
| Dependents of active duty | 0.591 | 0.353 | 0.990 | 0.0458 | —<sup>a</sup> |  |  |  |
| Retirees | 0.752 | 0.435 | 1.301 | 0.3084 | —<sup>a</sup> |  |  |  |
| Others | 1.056 | 0.752 | 1.483 | 0.7541 | —<sup>a</sup> |  |  |  |
| Service branch (ref: Air Force) |  |  |  |  |  |  |  |  |
| Army | 2.088 | 1.266 | 3.444 | 0.0039 | 1.582 | 0.930 | 2.691 | 0.0004 |
| Coast Guard | 0.564 | 0.187 | 1.702 | 0.3092 | 0.360 | 0.113 | 1.143 | 0.1017 |
| Other | 0.709 | 0.342 | 1.472 | 0.3565 | 0.686 | 0.328 | 1.437 | 0.7368 |
| Not applicable (non-active duty) | 1.039 | 0.697 | 1.550 | 0.8501 | 0.640 | 0.379 | 1.080 | 0.4056 |
| **Rank (ref: enlisted)** |  |  |  |  |  |  |  |  |
| Officer | 1.600 | 0.996 | 2.571 | 0.0518 | <sup>a</sup><br>— |  |  |  |
| Not applicable (non-active duty) | 1.016 | 0.721 | 1.431 | 0.9287 | <sup>a</sup><br>— |  |  |  |

<sup>a</sup>Beneficiary and rank are not included in the adjusted model due to collinearity with service branch.

**Table 5.**
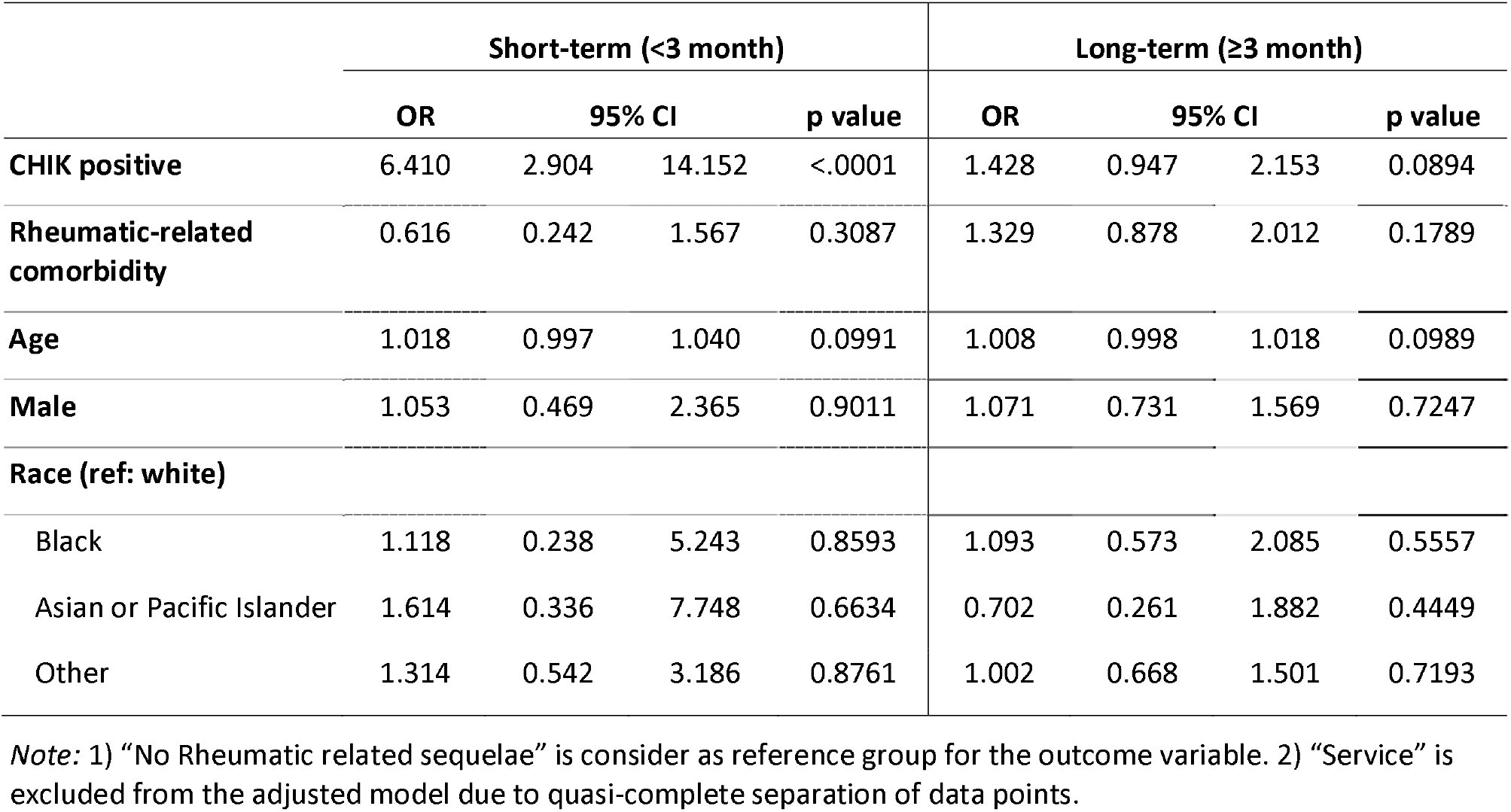
Adjusted odds ratio on rheumatic-related short-term and long-term sequelae (n = 975).

Among those with CHIKV infections, we found no association between post-CHIKV rheumatic sequelae and age, sex, race, active-duty status, or pre-existing comorbidities. Non-Army service branch and female sex was associated with a reduced odds of post-CHIKV rheumatological sequelae, but this was not statistically significant (Table 6).

**Table 6.**
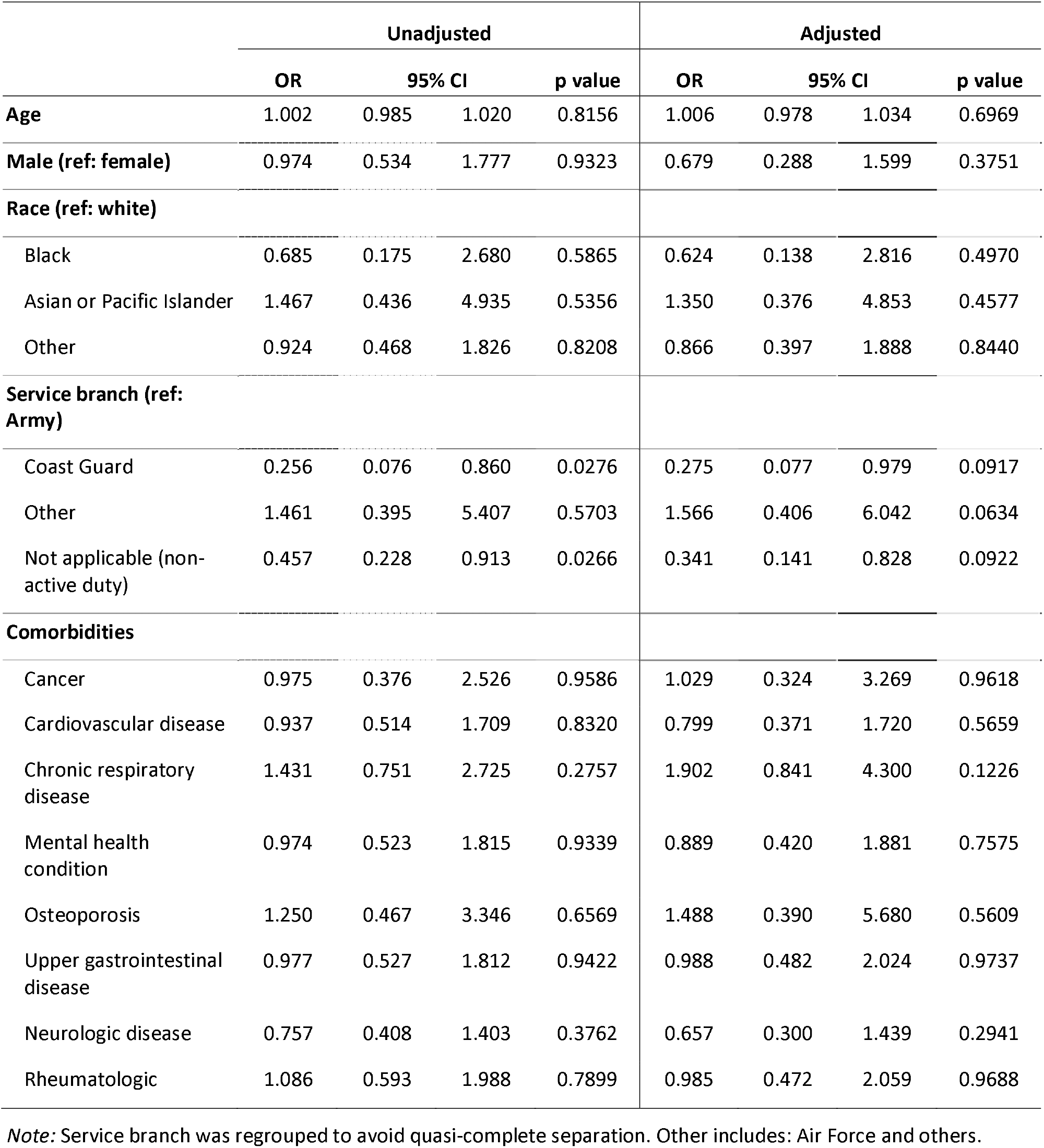
Unadjusted and adjusted odds ratio on rheumatic-related sequelae among people with CHIK infection (n = 195).

## Conclusion

Understanding the risk of CHIKV infection and complications in deploying U.S. military servicemembers and other MHS beneficiaries is important, particularly with upcoming possible CHIKV vaccine licensure. We found a very low number of CHIKV cases identified in the MHS, despite this study period covering the peak CHIKV epidemic in the Americas and leveraging data from about 9.5 million beneficiaries. Our findings correlate with low number of CHIKV infections in other U.S. MHS studies, although such studies were either smaller, earlier in the CHIKV epidemic and/or less comprehensive in their use of electronic medical record (eMR) data sources [28, 31]. Additionally, our findings correlate with a recent post-deployment serology study of 1500 deployments to South America and Puerto Rico with a low post-deployment seropositivity rate of 1.7% (highest in those serving in Puerto Rico) [32]. Our findings further agree with studies in other armed forces which also note a low frequency of CHIKV infections [28].

These low frequencies of infection may be attributed to effective personal mosquito avoidance or heterogeneity in risk exposure (for example, based on deployment role) when deployed or activated in CHIKV endemic countries or territories. Among CHIKV infections in servicemembers, we noted higher frequency Army service and enlisted ranks compared to matched servicemember controls without CHIKV; these groups have previously been noted at higher risk for other arboviruses [33, 34]. While the location of diagnosis may not reflect where the infection was acquired, the highest number of cases were diagnosed in Puerto Rico is consistent with the known high transmission of CHIKV during the neotropical epidemic [35].

While the overall number of infections was very low, we found a substantive risk of post-CHIKV rheumatological complications. We noted a considerable burden of disease with a median 7 healthcare encounters for this complication, and 25% of these complicated CHIKV cases required between 15 and 33 further encounters through the period of observation. These findings highlight that, while CHIKV infections are overall rare in MHS beneficiaries, patient morbidity in complicated CHIKV cases can be substantial, as noted in non-MHS studies [22].

The overall low number of CHIKV infections limited statistical power for examining risk factors for post-CHIKV sequelae. We found a non-statistically significant trend toward Army and male sex having a higher risk of rheumatic complications, but no other predictors were noted, including pre-existing comorbidities. Further study of cumulative CHIKV infections in MHS-based virtual cohorts over time may identify clearer risk factors for this complication, and this may assist in more precise prognostication of CHIKV cases in the MHS. As this study comprises the largest number of CHIKV infections in any military study, we would conclude that prediction of MHS CHIKV cases may progress to complicated rheumatological sequelae remains challenging, underscoring the need for optimal prevention of CHIKV infection.

There were several strengths to this study. The use of the MHS includes a substantive number of beneficiaries who deploy or are stationed in CHIKV endemic regions and allowed for long term follow-up of cases to determine longer term complications. We derived CHIKV cases from a range of eMR data sources, including clinical microbiology laboratory results, to identify more infections than prior MHS-based studies [28–31]. Our study limitations included the use of ICD codes to diagnose CHIKV and post-CHIKV complications; prior evaluations of CHIKV ICD diagnosis coding have noted imperfect specificity [31] and this study was unable to perform chart adjudications. Conversely, CHIKV diagnoses may have been missed in some clinical presentations, and not all cases may have presented to care (including during deployments).

Future directions include further studies which could also examine for non-rheumatological CHIKV sequelae such as neurocognitive complications, mood, and long-term functional impact (for example, [36, 37]). Finally, studies could leverage sera routinely collected from the U.S. military servicemembers and used for arboviral research (i.e., the Department of Defense Sera Repository [32–34]) to identify prognostic and treatment related biomarkers for those beneficiaries with persistent post-CHIKV sequelae, as well as enabling ongoing serosurveillance of the risk and risk factors of CHIKV in deployed military, particularly during an era of increasing CHIKV risk in the Americas [9].

## Data Availability

Data for this study are available from the Infectious Disease Clinical Research Program (IDCRP), headquartered at the Uniformed Services University of the Health Sciences (USU), Department of Preventive Medicine and Biostatistics. Review by the USU Institutional Review Board is required for use of the data used in this protocol. Furthermore, the data set includes Military Health System data collected under a Data Use Agreement that requires accounting for uses of the data. Data requests may be sent to: Address: 6270A Rockledge Drive, Suite 250, Bethesda, MD 20817.

## Disclaimer

The contents of this publication are the sole responsibility of the author(s) and do not necessarily reflect the views, opinions, or policies of Uniformed Services University of the Health Sciences; the Department of Defense; the Departments of the Army, Navy, or Air Force; the Defense Health Agency; the National Institutes of Health, the Department of Health and Human Services or the Henry M. Jackson Foundation for the Advancement of Military Medicine, Inc. Mention of trade names, commercial products, or organizations does not imply endorsement by the U.S. government. The investigators have adhered to the policies for protection of human subjects as prescribed in 45 CFR 46.

## Funding statement

The protocol was executed by the Infectious Disease Clinical Research Program (IDCRP), a Department of Defense program executed by the Uniformed Services University of the Health Sciences through a cooperative agreement by the Henry M. Jackson Foundation for the Advancement of Military Medicine, Inc.. This project has been funded in part by the National Institute of Allergy and Infectious Diseases at the National Institutes of Health, under an interagency agreement (Y1-AI-5072); and the Military Infectious Diseases Research Program. The authors declare no conflicts of interest.

## Supporting information

**S1 Fig. Chikungunya virus case identification from U.S. Military Health System electronic medical record systems.** TEDNI–TRICARE Encounter Data Non-Institutional; CAPER–Comprehensive Ambulatory/Professional Encounter Record.

**S1 Table. International Classification of Diseases, 9th Revision (ICD-9) and ICD-10 codes (including cross-walked ICD-9 codes) used to identify rheumatological healthcare encounters in cases and controls.**

**S2 Table. Long description of rheumatological healthcare encounters by chikungunya virus (CHIKV) infection status.**

## Notes

### Competing Interest Statement

The authors have declared no competing interest.

