## Supplemental Materials for "The risk and risk factors of chikungunya virus infection and rheumatological sequelae in a cohort of U.S. Military Health System beneficiaries: implications for the vaccine era"

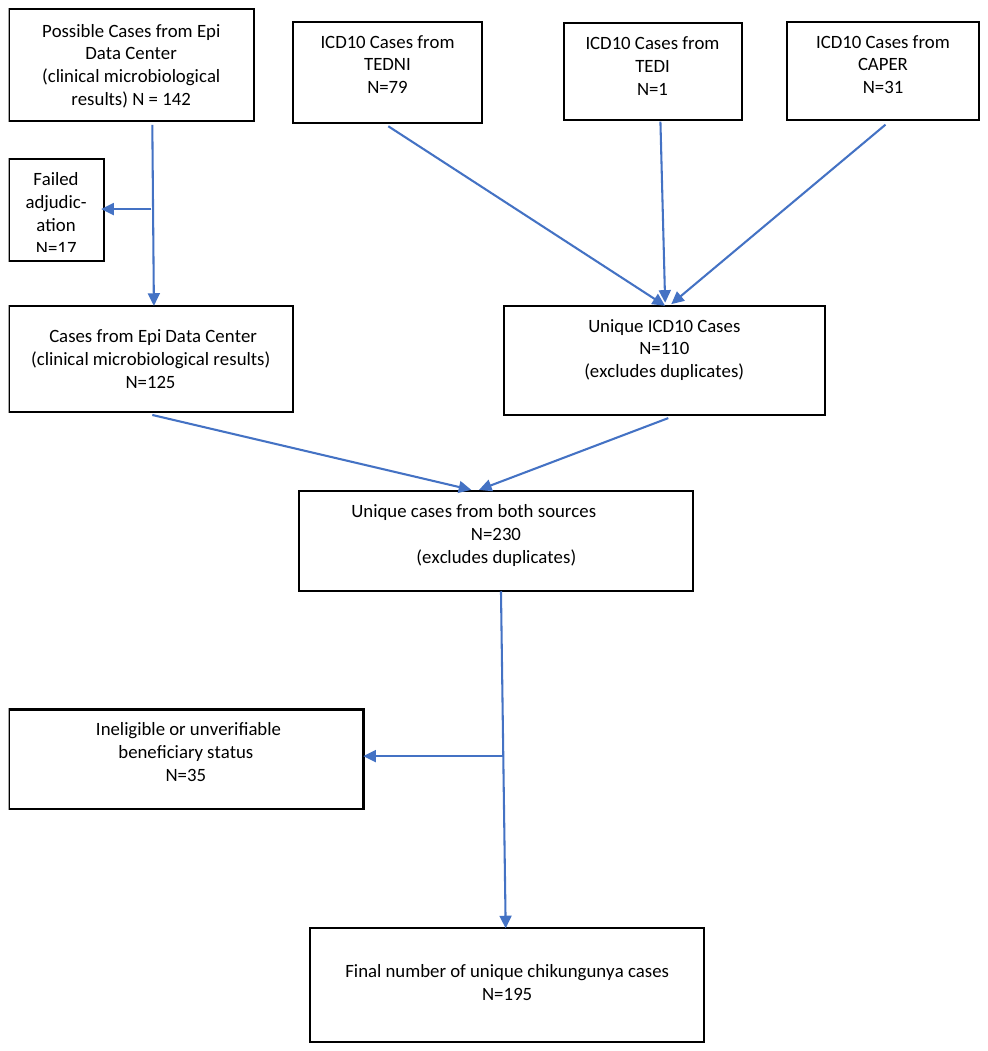

**Figure S1.** CHIKV case identification from MHS EMR systems. TEDNI - TRICARE Encounter Data Non-Institutional; CAPER - Comprehensive Ambulatory/Professional Encounter Record

| **Table S1. ICD9 and ICD10 codes (including cross-walked ICD9 codes) used to identify rheumatological healthcare encounters in cases and controls** | |
| --- | --- |
| ICD9 | DESCRIPTION |
| 71150 | ARTHROPATHY SITE UNSPECIFIED ASSOCIATED WITH OTHER VIRAL DISEASES |
| 71151 | ARTHROPATHY INVOLVING SHOULDER REGION ASSOCIATED WITH OTHER VIRAL DISEASES |
| 71152 | ARTHROPATHY INVOLVING UPPER ARM ASSOCIATED WITH OTHER VIRAL DISEASES |
| 71153 | ARTHROPATHY INVOLVING FOREARM ASSOCIATED WITH OTHER VIRAL DISEASES |
| 71154 | ARTHROPATHY INVOLVING HAND ASSOCIATED WITH OTHER VIRAL DISEASES |
| 71155 | ARTHROPATHY INVOLVING PELVIC REGION AND THIGH ASSOCIATED WITH OTHER VIRAL DISEASES |
| 71156 | ARTHROPATHY INVOLVING LOWER LEG ASSOCIATED WITH OTHER VIRAL DISEASES |
| 71157 | ARTHROPATHY INVOLVING ANKLE AND FOOT ASSOCIATED WITH OTHER VIRAL DISEASES |
| 71158 | ARTHROPATHY INVOLVING OTHER SPECIFIED SITES ASSOCIATED WITH OTHER VIRAL DISEASES |
| 71159 | ARTHROPATHY INVOLVING MULTIPLE SITES ASSOCIATED WITH OTHER VIRAL DISEASES |
| 7136 | ARTHROPATHY ASSOCIATED WITH HYPERSENSITIVITY REACTION |
| 7136 | ARTHROPATHY ASSOCIATED WITH HYPERSENSITIVITY REACTION |
| 7137 | OTHER GENERAL DISEASES WITH ARTICULAR INVOLVEMENT |
| 7138 | ARTHROPATHY ASSOCIATED WITH OTHER CONDITIONS CLASSIFIABLE ELSEWHERE |
| 7140 | RHEUMATOID ARTHRITIS |
| 71489 | OTHER SPECIFIED INFLAMMATORY POLYARTHROPATHIES |
| 7149 | UNSPECIFIED INFLAMMATORY POLYARTHROPATHY |
| 71500 | OSTEOARTHROSIS GENERALIZED INVOLVING UNSPECIFIED SITE |
| 71500 | OSTEOARTHROSIS GENERALIZED INVOLVING UNSPECIFIED SITE |
| 71509 | OSTEOARTHROSIS GENERALIZED INVOLVING MULTIPLE SITES |
| 71580 | OSTEOARTHROSIS INVOLVING OR WITH MORE THAN ONE SITE BUT NOT SPECIFIED AS GENERALIZED AND INVOLVING UNSPECIFIED SITE |
| 71589 | OSTEOARTHROSIS INVOLVING OR WITH MULTIPLE SITES BUT NOT SPECIFIED AS GENERALIZED |
| 71589 | OSTEOARTHROSIS INVOLVING OR WITH MULTIPLE SITES BUT NOT SPECIFIED AS GENERALIZED |
| 71649 | TRANSIENT ARTHROPATHY INVOLVING MULTIPLE SITES |
| 71659 | UNSPECIFIED POLYARTHROPATHY OR POLYARTHRITIS INVOLVING MULTIPLE SITES |
| 71689 | OTHER SPECIFIED ARTHROPATHY INVOLVING MULTIPLE SITES |
| 71699 | UNSPECIFIED ARTHROPATHY INVOLVING MULTIPLE SITES |
| 71949 | PAIN IN JOINT INVOLVING MULTIPLE SITES |
| 71959 | STIFFNESS OF JOINT NOT ELSEWHERE CLASSIFIED INVOLVING MULTIPLE SITES |
| 71969 | OTHER SYMPTOMS REFERABLE TO JOINT OF MULTIPLE SITES |
| 71989 | OTHER SPECIFIED DISORDERS OF JOINT OF MULTIPLE SITES |
| 71989 | OTHER SPECIFIED DISORDERS OF JOINT OF MULTIPLE SITES |
| 71999 | UNSPECIFIED JOINT DISORDER OF MULTIPLE SITES |
| 725 | POLYMYALGIA RHEUMATICA |
| 7290 | RHEUMATISM UNSPECIFIED AND FIBROSITIS |
| 7291 | MYALGIA AND MYOSITIS, UNSPECIFIED |
| 78079 | OTHER MALAISE AND FATIGUE |
| 78079 | OTHER MALAISE AND FATIGUE |
| 78079 | OTHER MALAISE AND FATIGUE |
| 78079 | OTHER MALAISE AND FATIGUE |
| 78079 | OTHER MALAISE AND FATIGUE |
| V821 | SCREENING FOR RHEUMATOID ARTHRITIS |
| V822 | SCREENING FOR OTHER RHEUMATIC DISORDERS |
| ICD10 | DESCRIPTION |
| M01X0 | DIRECT INFECTION OF UNSPECIFIED JOINT IN INFECTIOUS AND PARASITIC DISEASES CLASSIFIED ELSEWHERE |
| M01X19 | DIRECT INFECTION OF UNSPECIFIED SHOULDER IN INFECTIOUS AND PARASITIC DISEASES CLASSIFIED ELSEWHERE |
| M01X29 | DIRECT INFECTION OF UNSPECIFIED ELBOW IN INFECTIOUS AND PARASITIC DISEASES CLASSIFIED ELSEWHERE |
| M01X39 | DIRECT INFECTION OF UNSPECIFIED WRIST IN INFECTIOUS AND PARASITIC DISEASES CLASSIFIED ELSEWHERE |
| M01X49 | DIRECT INFECTION OF UNSPECIFIED HAND IN INFECTIOUS AND PARASITIC DISEASES CLASSIFIED ELSEWHERE |
| M01X59 | DIRECT INFECTION OF UNSPECIFIED HIP IN INFECTIOUS AND PARASITIC DISEASES CLASSIFIED ELSEWHERE |
| M01X69 | DIRECT INFECTION OF UNSPECIFIED KNEE IN INFECTIOUS AND PARASITIC DISEASES CLASSIFIED ELSEWHERE |
| M01X79 | DIRECT INFECTION OF UNSPECIFIED ANKLE AND FOOT IN INFECTIOUS AND PARASITIC DISEASES CLASSIFIED ELSEWHERE |
| M01X8 | DIRECT INFECTION OF VERTEBRAE IN INFECTIOUS AND PARASITIC DISEASES CLASSIFIED ELSEWHERE |
| M01X9 | DIRECT INFECTION OF MULTIPLE JOINTS IN INFECTIOUS AND PARASITIC DISEASES CLASSIFIED ELSEWHERE |
| M0220 | POSTIMMUNIZATION ARTHROPATHY, UNSPECIFIED SITE |
| M364 | ARTHROPATHY IN HYPERSENSITIVITY REACTIONS CLASSIFIED ELSEWHERE |
| M029 | REACTIVE ARTHROPATHY, UNSPECIFIED |
| M1480 | ARTHROPATHIES IN OTHER SPECIFIED DISEASES CLASSIFIED ELSEWHERE, UNSPECIFIED SITE |
| M069 | RHEUMATOID ARTHRITIS, UNSPECIFIED |
| M064 | INFLAMMATORY POLYARTHROPATHY |
| M064 | INFLAMMATORY POLYARTHROPATHY |
| M150 | PRIMARY GENERALIZED (OSTEO)ARTHRITIS |
| M159 | POLYOSTEOARTHRITIS, UNSPECIFIED |
| M150 | PRIMARY GENERALIZED (OSTEO)ARTHRITIS |
| M158 | OTHER POLYOSTEOARTHRITIS |
| M153 | SECONDARY MULTIPLE ARTHRITIS |
| M158 | OTHER POLYOSTEOARTHRITIS |
| M1289 | OTHER SPECIFIC ARTHROPATHIES, NOT ELSEWHERE CLASSIFIED, MULTIPLE SITES |
| M130 | POLYARTHRITIS, UNSPECIFIED |
| M1289 | OTHER SPECIFIC ARTHROPATHIES, NOT ELSEWHERE CLASSIFIED, MULTIPLE SITES |
| M129 | ARTHROPATHY, UNSPECIFIED |
| M2550 | PAIN IN UNSPECIFIED JOINT |
| M2560 | STIFFNESS OF UNSPECIFIED JOINT, NOT ELSEWHERE CLASSIFIED |
| R29898 | OTHER SYMPTOMS AND SIGNS INVOLVING THE MUSCULOSKELETAL SYSTEM |
| M2510 | FISTULA, UNSPECIFIED JOINT |
| M2580 | OTHER SPECIFIED JOINT DISORDERS, UNSPECIFIED JOINT |
| M259 | JOINT DISORDER, UNSPECIFIED |
| M353 | POLYMYALGIA RHEUMATICA |
| M790 | RHEUMATISM, UNSPECIFIED |
| M609 | MYOSITIS, UNSPECIFIED |
| M791 | MYALGIA |
| M797 | FIBROMYALGIA |
| [R53.8](https://www.icd10data.com/ICD10CM/Codes/R00-R99/R50-R69/R53-/R53.8) | OTHER MALAISE AND FATIGUE |
| Z13828 | ENCOUNTER FOR SCREENING FOR OTHER MUSCULOSKELETAL DISORDER |
| Z1389 | ENCOUNTER FOR SCREENING FOR OTHER DISORDER |

| **Table S2. Long description of rheumatological healthcare encounters by CHIKV infection status** | | | | | | |
| --- | --- | --- | --- | --- | --- | --- |
|  | | **CHIKV** | | | | **ALL** |
|  |  | **Negative** | | **Positive** | |  |
|  |  | **N** | **%** | **N** | **%** | **N** |
| **Disorder** | **Long Description** |  |  |  |  |  |
| **RHEUMATIC DISORDER** |  |  |  |  |  |  |
| **INFECTIVE ARTHRITIS AND OSTEOMYELITIS** | DIRECT INFECTION OF UNSPECIFIED JOINT IN INFECTIOUS AND PARASITIC DISEASES CLASSIFIED ELSEWHERE | . | . | 3 | 100.00 | 3 |
| **MALAISE AND FATIGUE** | OTHER FATIGUE | 93 | 85.32 | 16 | 14.68 | 109 |
|  | OTHER MALAISE | 15 | 78.95 | 4 | 21.05 | 19 |
|  | OTHER MALAISE AND FATIGUE | 11 | 84.62 | 2 | 15.38 | 13 |
|  | WEAKNESS | 47 | 79.66 | 12 | 20.34 | 59 |
| **OSTEOARTHRITIS** | OSTEOARTHROSIS, GENERALIZED, INVOLVING MULTIPLE SITES | 1 | 33.33 | 2 | 66.67 | 3 |
|  | OTHER POLYOSTEOARTHRITIS | . | . | 4 | 100.00 | 4 |
|  | PRIMARY GENERALIZED (OSTEO)ARTHRITIS | 3 | 13.64 | 19 | 86.36 | 22 |
| **OTHER CONNECTIVE TISSUE DISEASE** | OTHER SYMPTOMS AND SIGNS INVOLVING THE MUSCULOSKELETAL SYSTEM | 47 | 54.65 | 39 | 45.35 | 86 |
|  | POLYMYALGIA RHEUMATICA | . | . | 1 | 100.00 | 1 |
|  | RHEUMATISM, UNSPECIFIED | 1 | 100.00 | . | . | 1 |
| **OTHER NON-TRAUMATIC JOINT DISORDERS** | ARTHROPATHY, UNSPECIFIED | 6 | 46.15 | 7 | 53.85 | 13 |
|  | JOINT DISORDER, UNSPECIFIED | 2 | 100.00 | . | . | 2 |
|  | PAIN IN JOINT INVOLVING MULTIPLE SITES | 1 | 7.14 | 13 | 92.86 | 14 |
|  | PAIN IN UNSPECIFIED JOINT | 20 | 41.67 | 28 | 58.33 | 48 |
|  | POLYARTHRITIS, UNSPECIFIED | 3 | 42.86 | 4 | 57.14 | 7 |
|  | STIFFNESS OF UNSPECIFIED JOINT, NOT ELSEWHERE CLASSIFIED | 15 | 88.24 | 2 | 11.76 | 17 |
| **OTHER SCREENING FOR SUSPECTED CONDITIONS (NOT MENTAL DISORDERS OR INFECTIOUS DISEASE)** | ENCOUNTER FOR SCREENING FOR OTHER DISORDER | 179 | 74.27 | 62 | 25.73 | 241 |
|  | ENCOUNTER FOR SCREENING FOR OTHER MUSCULOSKELETAL DISORDER | 1 | 100.00 | . | . | 1 |
| **RHEUMATOID ARTHRITIS AND RELATED DISEASE** | INFLAMMATORY POLYARTHROPATHY | . | . | 2 | 100.00 | 2 |
|  | RHEUMATOID ARTHRITIS | . | . | 8 | 100.00 | 8 |
|  | RHEUMATOID ARTHRITIS, UNSPECIFIED | 3 | 21.43 | 11 | 78.57 | 14 |
